# A Statistical Framework to Identify Cell Types Whose Genetically Regulated Proportions are Associated with Complex Diseases

**DOI:** 10.1101/2021.02.25.21252462

**Authors:** Wei Liu, Wenxuan Deng, Ming Chen, Zihan Dong, Biqing Zhu, Zhaolong Yu, Daiwei Tang, Maor Sauler, Louise V. Wain, Michael H. Cho, Naftali Kaminski, Hongyu Zhao

**Affiliations:** Program of Computational Biology and Bioinformatics, Yale University, New Haven, CT, USA 06510; Department of Biostatistics, Yale School of Public Health, New Haven, CT, USA 06510; Pulmonary, Critical Care and Sleep Medicine, Yale School of Medicine, New Haven, CT, USA 06510; Department of Health Sciences, University of Leicester, Leicester, United Kingdom; National Institute for Health Research, Leicester Respiratory Biomedical Research Centre, Glenfield Hospital, Leicester, United Kingdom; Channing Division of Network Medicine, Brigham and Women’s Hospital, Harvard Medical School, Boston, MA; Pulmonary and Critical Care Medicine, Brigham and Women’s Hospital, Harvard Medical School, Boston, MA.

**Keywords:** GWAS, cell type proportion imputation, single cell data, complex traits

## Abstract

Finding disease-relevant tissues and cell types can facilitate the identification and investigation of functional genes and variants. In particular, cell type proportions can serve as potential disease predictive biomarkers. Here, we introduce a novel statistical framework, cell-type Wide Association Study (cWAS), that integrates genetic data with transcriptomics data to identify cell types whose genetically regulated proportions (GRPs) are disease/trait-associated. On simulated and real GWAS data, cWAS showed substantial statistical power with newly identified significant GRP associations in disease-associated tissues. More specifically, GRPs of endothelial and myofibroblasts in lung tissue were associated with Idiopathic Pulmonary Fibrosis and Chronic Obstructive Pulmonary Disease, respectively. For breast cancer, the GRP of blood CD8^+^ T cells was negatively associated with breast cancer (BC) risk as well as survival. Overall, cWAS is a powerful tool to reveal cell types associated with complex diseases mediated by GRPs.

## Introduction

Despite the great success of genome-wide association studies (GWAS), it has been challenging to identify disease-causing genes and variants. To better design functional studies of GWAS implicated SNPs, it is important to identify tissues and cell types most relevant to a disease. Several statistical approaches have been developed for this purpose^1–3^. In general, these methods aim to detect statistically significant overlap between GWAS signals and annotated functional regions in specific tissues and cell types, where the annotated functional regions are curated from other data sources, such as ENCODE and Roadmap Epigenomics data and single cell data. Although such analyses have led to novel insights on disease mechanisms^1,4–7^, the cell types associated with the majority of genomic regions remain to be discovered.

Several studies have found that the proportions of cell types are not only associated with disease incidence^8,9^ but also disease prognosis^10,11^. Single cell RNA-seq (scRNA-seq) technologies have been used to identify cell type proportions that impact human diseases and traits^12^. However, several intrinsic characteristics of single cell data make disease-cell type proportion association analysis challenging. First, high expense and technical noise (e.g., high sparsity of gene expression) limit the number of samples analyzed and quality of cell type composition estimation, leading to low power in association analysis. Second, cell type compositions measured in single cell experiments are highly dependent on the biopsy samples and do not necessarily reflect the true cell type compositions in the corresponding tissue^13^. Instead of directly calculating cell type proportions from scRNA-seq data, cell type proportions can also be inferred through deconvolution of bulk RNA-sequencing (RNA-seq) data available with larger sample sizes. Many computational methods have been developed to estimate cell type proportions in bulk RNA-seq data using cell type-specific gene expression signatures derived from either microarray or scRNA-seq reference^12^. Compared with biopsy samples in single cell analyses, tissue samples for bulk analysis might better represent the original cell type compositions^8,12^.

For both single cell and bulk data, cell type proportions can be affected by various factors including disease status and treatment effects. Consequently, the observed cell type proportion differences between disease and healthy individuals might be the outcome of the disease and environmental factors instead of disease causes.

Unlike assayed gene expression levels, genotypes are less likely to be affected by confounding factors and reverse causation. The same idea underlies Mendelian randomization methods to infer causal factors for different traits^9,10,14^. In this paper, we examined genetically regulated proportions (GRPs) of cell types. We note that cell type proportions are heritable^11,15^, suggesting the feasibility of inferring cell type proportions based on genotypes. Cell type proportions can vary substantially in patients with different diseases^16^. We introduce a new framework, cell type Wide Association Study (cWAS), to consider the GRPs of cell types as contributors to human disease. Through simulation studies and real data analyses across 55 traits in 36 tissues, cWAS showed higher statistical power in identifying disease-cell type proportion associations than typical cell-disease association identification approaches like FUMA^3^. In summary, cWAS offers a novel way to understand human diseases in a cell-type specific manner.

## Results

### Model summary

We propose a statistical framework to identify cell types whose GRPs are associated with diseases. The framework consists of two parts (**Figure 1**). First, under the assumption that there exist signature genes signifying specific cell types (consistent with previous methods^15,17^), we infer GRPs of cell types through deconvolution of the imputed tissue-specific gene expression levels based on cis-SNP genotypes from eQTL data. Second, we combine the GRPs with disease phenotype information to identify cell-type proportion associations with disease phenotypes.

**Figure 1.**
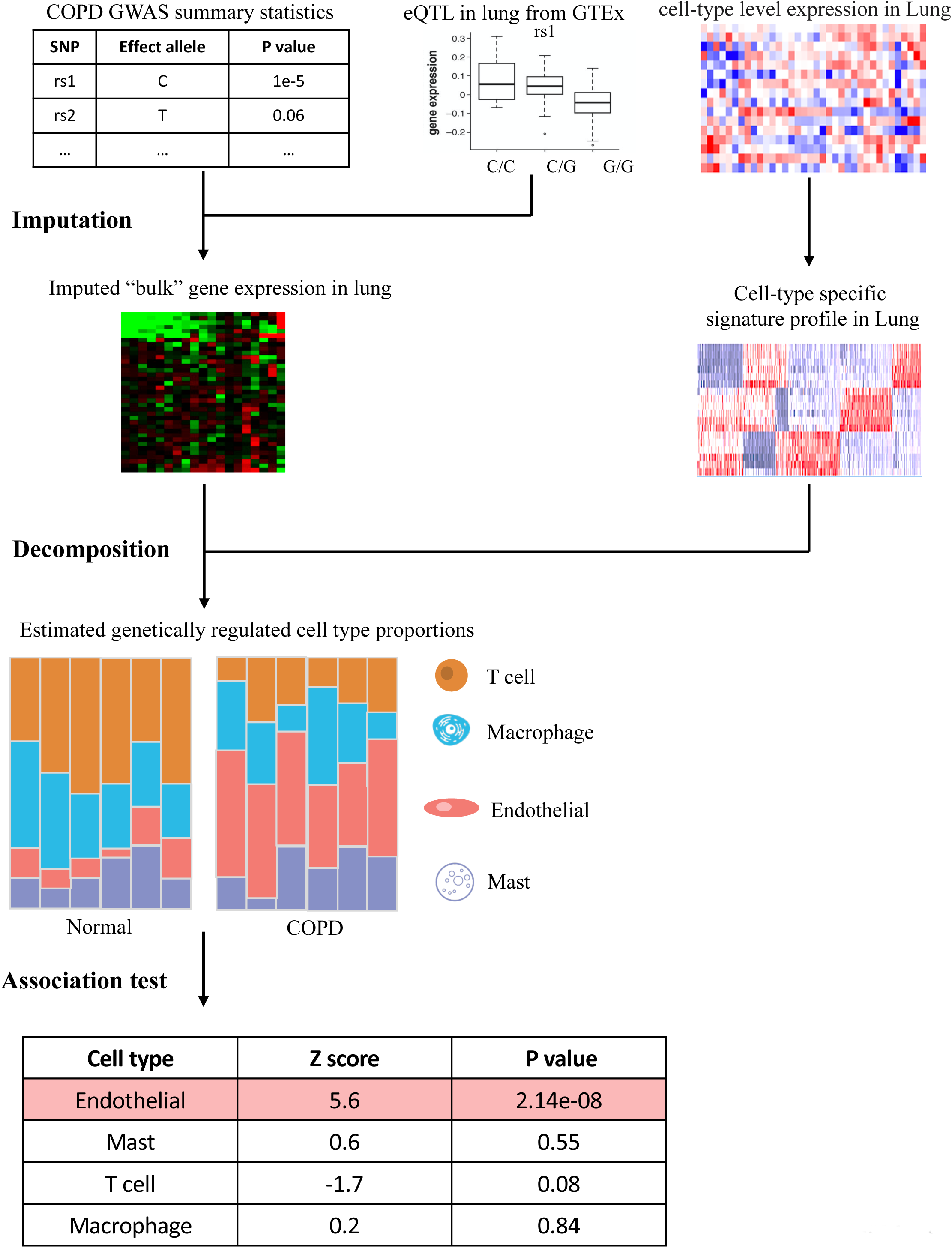
The schematic framework of cWAS. Bulk gene expression levels are firstly imputed based on each individual’s genotypes. Combined with a signature gene expression matrix for different cell types, imputed gene expression data for each tissue are used to infer cell type compositions. Comparing different genetically inferred cell type compositions in case and control groups, cWAS can identify cell types whose genetic-regulated proportions are associated with a trait of interest.

In the first step, we build tissue-specific gene expression imputation models using the elastic net, similar to previous Transcriptome-wide association study (TWAS) methods^18–20^. With the imputation weights 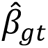, we obtain the estimation of genetically regulated tissue-level gene expression for gene *g* in tissue *t* as 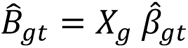, where *X_g_* is the genotype matrix of cis-SNPs around gene *g*. With pre-obtained cell-type specific gene expression levels for signature genes, we deconvolute the genetically imputed tissue-level expression data through the following model:

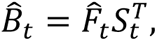

where *S_t_* ∈ *R^G^*^×*C*^ is the cell-type specific gene expression level matrix in tissue *t* for *G* signature genes across *C* cell types, 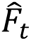 is the estimated GRPs for all cell types in tissue *t*, and 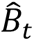 is the imputed gene expression level matrix for all signature genes in tissue *t*. For a specific cell type *c*, we assess its GRP association with phenotype *Y* using the following model:

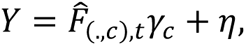

where *γ_c_* is the effect of GRP for cell type *c* on the trait and η is noise. 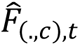 is the estimated GRPs of a cell type *c*, which is the *c*th column of the 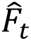 matrix. However, individual-level genotype data are not always available for GWAS, which makes the direct estimation of *γ_c_* from the above two-step procedure unfeasible. With only summary statistics available, we propose to use the following approach to assessing the association between GRPs of a cell type *c* and traits

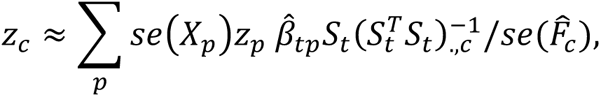

where *se*(*X_p_*) is the genotype standard deviation of SNP *p*, calculated from a reference panel; *z_p_* is the GWAS z score for SNP *p*; 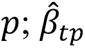, is the imputed tissue-level gene expression vector of SNP *p* across *G* signature genes in tissue *t*, and (.)*_.,c_* stands for the *c*th column vector of the corresponding matrix. cWAS takes the GWAS summary statistics as the input, which provides an indirect way of estimating cell-type GRP associations with diseases that do not require individual-level data. More model details are presented in the methods section, and the cWAS framework for GWAS summary statistics is available at https://github.com/vivid-/cWAS.

### Simulation studies

To evaluate cWAS performance in identifying cell type proportions associated with a disease, we considered several simulation settings (**Methods**). We simulated disease phenotypes based on genetically predicted proportions of M1 macrophages in whole blood, using 10,000 individuals randomly sampled from UK Biobank^21^. Under moderate heritability settings, where genetic-regulated cell type proportions explain 1% to 9% of the phenotype variances, cWAS had at least 98% power to identify M1 macrophages’ association with the phenotype when all signature genes were known and used (**Figure 2a**, the purple dashed line). Furthermore, M1 macrophage was identified as the most significant cell type in at least 70% of 600 replicates (**Figure 2b**) when heritability was 4% or higher, and the effect of M1 macrophages identified by cWAS had the same direction as that simulated in at least 90% of 600 replicates, while FUMA only identified macrophages as the significant cell type in around 15% of 600 replicates (**S Table 1**). When we simulated phenotypes independent of cell type proportions in whole blood tissue, cWAS had a well-controlled type I error rate (**Figure 2c**).

**Figure 2.**
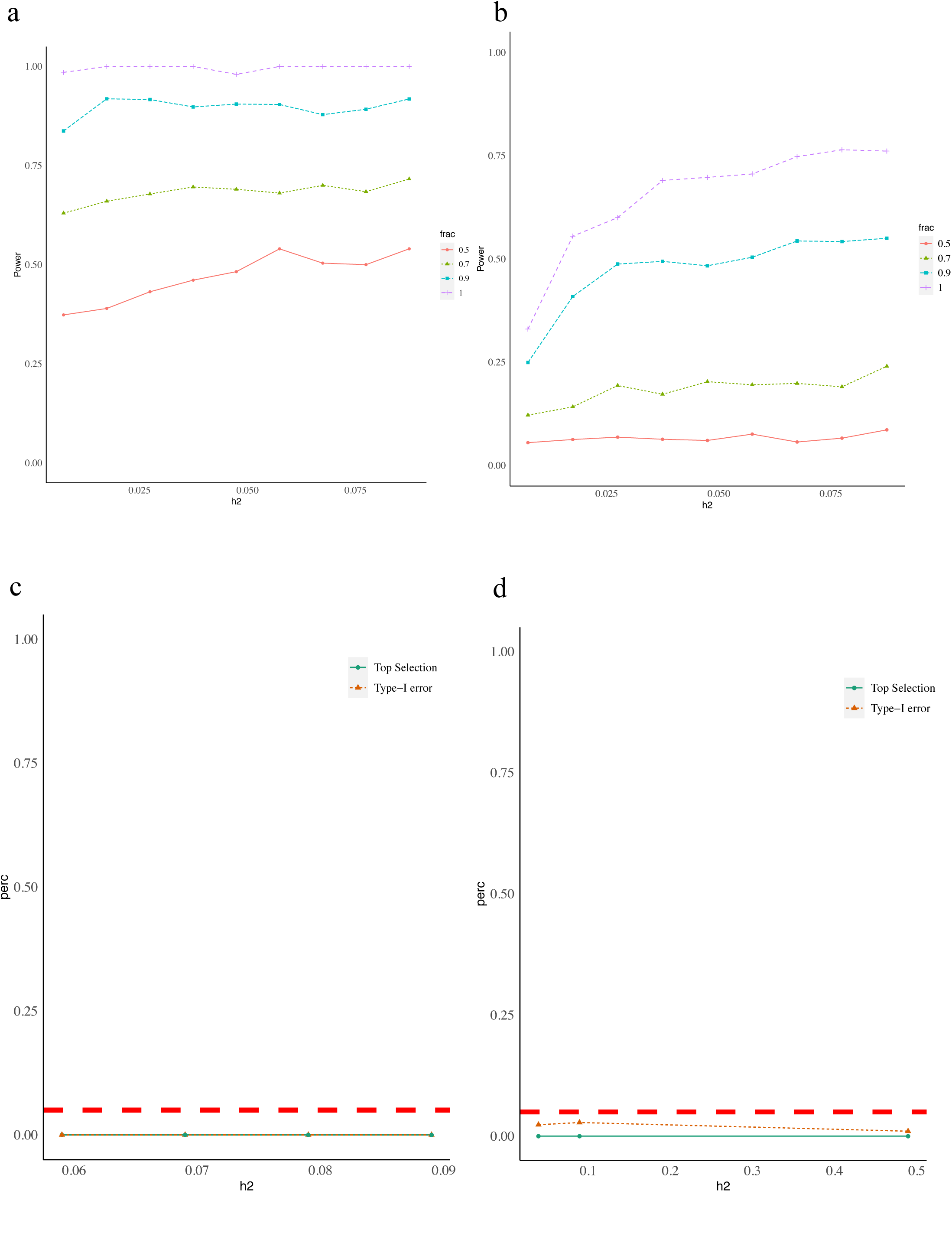
High power of cWAS in simulation studies with a controlled type I error rate. Different colors indicate different proportions (0.5, 0.7, 0.9, and 1) of signature genes used in the cWAS test. The phenotypic variance explained by the genetically regulated cell type proportions (M1 macrophages) ranges from 0.01 to 0.09 for panels A and B, respectively. a) Each line represents the percentage of simulations where cWAS identified the M1 macrophages as associated with simulated phenotypes. b) This figure shows the proportion of times that the M1 macrophage was identified as the most significant cell type whose proportion was associated with simulated phenotypes. The line represents the rate (top selection rate) under settings with different proportions of known signature genes. For panels c and d, we simulated phenotypes based on the genetic-regulated proportion of basal cells in lung tissue with heritability being 0.05, 0.1, or 0.5. c) All signature genes in whole blood are known when conducting the cWAS test. The red dashed line indicates the 5% type I error. The green line indicates the proportion of simulations where any cell type in whole blood was selected as associated with the simulated disease status, the orange line indicates the proportion of simulations where M1 macrophages were selected as associated with the simulated disease. d) Only 50% of signature genes in whole blood are known.

One critical point of cWAS is the reliability of cell type specific gene expression signatures. Many cell-type deconvolution methods also depend on the accurate curation of the signature matrix, such as those from microarray data of known cell types (like the LM22 matrix used in CIBERSORT^17^). However, in many cases, we have to derive a signature matrix from single-cell data, which are usually highly sparse and only include cell type-specific expression levels of a subset of signature genes. Consequently, the signature genes curated from single-cell data may be incomplete compared to those from more informative data sources, such as RNA-seq assayed in known cell types. To evaluate the impact of incomplete genes in the signature matrix, we considered using a subset (50%-90%) of signature genes in cWAS. When only half of the signature genes were used, there was a significant drop in statistical power although the type I error was still well-controlled (**Figure 2d**). With an increasing proportion of signature genes used, there was improved power in identifying associated cell types (**Figure 2a**).

### Trait-tissue association patterns

To further study disease-cell type proportion associations, we applied cWAS to GWAS summary data from 55 traits (**S Table 2**, including autoimmune diseases, psychiatric disorders, and other traits like lipids and height) together with scRNA-seq data from the Human Cell Landscape (HCL)^22^. We identified trait-associated cell types in 23 adult non-brain tissues and 13 fetal brain tissues (**S Table 3**) using eQTLs for curated signature genes (**Methods, S Fig. 1**). Consistent with findings from other methods, we found that the most significant cell types are usually present in the trait-associated tissues^1,23^ (**Figure 3a, S Fig. 2**) supporting the validity of cWAS, e.g., oligodendrocytes from fetal brain amygdala for autism spectrum disorder (ASD) (p= 3.0e-3), myeloid progenitor cells from whole blood for Crohn’s disease (p=3.6e-5), and endothelial cells from a tibial artery for heart rate (HR) (p=4.0e-9). Several traits showed global cell type proportion associations across multiple tissues, e.g., height and body mass index (BMI). This can be partly explained by large sample sizes in BMI and height GWAS, as we also observed a significant positive correlation (p=8.4e-4, cor=0.88) between the number of associated cell types and the sample size of BMI and height GWAS when we down-sampled the GWAS results (**Methods**). Notably, cWAS identified many cell type-trait associations in unexpected tissues. Many of them are immune cells, for example, neutrophil cells in fetal brain frontal cortex are associated with systemic lupus erythematosus (SLE) (p=5.8e-4), and macrophages from subcutaneous adipose and neutrophils from the left ventricle of the heart are associated with anxiety disorders (ADIS) (p=7.4e-3 and 1.6e-3, respectively).

**Figure 3.**
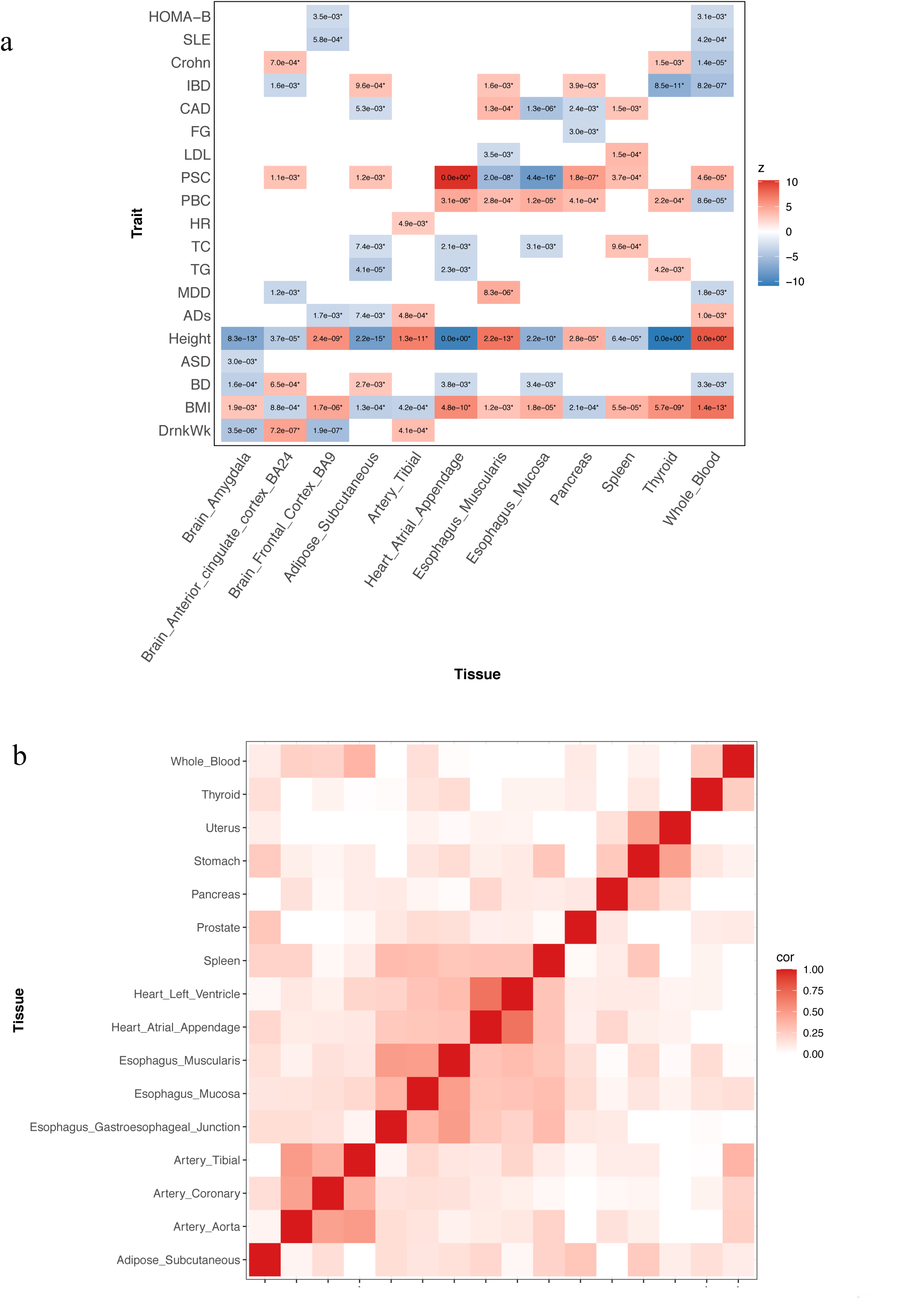
Disease-cell type associations in multiple tissues. a) Across 12 tissues, the z scores derived from the test statistics quantify the associations between genetically regulated cell type proportions and diseases. If there is no cell type significantly associated with a disease after Bonferroni correction, the corresponding entry is blank. The number in each block indicates the p-value of the most significant association between the cell type proportion of the corresponding tissue and the disease. HOMA-B: beta-cell function; SLE: Systemic Lupus Erythematosus; Crohn: Crohn’s disease; IBD: Inflammatory Bowel Disease; CAD: Coronary Artery Disease; FG: Fasting Glucose; LDL: LDL cholesterol; PSC: Primary Sclerosing Cholangitis; PBC: Primary Biliary Cirrhosis; HR: Resting Heart Rate; TC: Total Cholesterol; TG: Triglycerides; MDD: Major Depressive Disorder; ADs: Anxiety Disorder; Height: Height; ASD: Autism Spectrum Disorder; BD: Bipolar Disorder; BMI: Body Mass Index; DrnkWk: Drinks per Week. b) For any tissue pair, we only considered shared cell types and treated their proportion associations across 55 tissues. Tissue-tissue correlations were calculated based on the cell type-disease associations for the shared cell types. The darker color indicates a higher significance level.

Since several cell types (**S Table 4**), especially immune cells, are present in multiple adult tissues, we further investigated whether those identified disease-associated immune cell types above are due to true biological process or false positives by studying tissue-tissue correlations based on shared cell types’ associations with traits (**Methods**). Compared to biologically unrelated tissue pairs, the results showed a higher correlation among tissues with similar biological functions (**Figure 3b**), such as artery tissues (tibial artery, coronary artery, and aorta artery), heart tissues (left heart ventricle and heart atrial appendage), and esophagus tissues (esophagus muscularis and esophagus mucosa). This finding suggests that cell types are more likely to be identified as trait-associated in disease-related tissues even though the same cell types may exist in multiple tissues.

We also evaluated correlations among traits based on their associations with different cell types across 23 adult non-brain tissues and 13 fetal brain tissues, respectively (**S Table 5**). In 23 adult non-brain tissues, we identified high correlations among many traits, e.g., autoimmune diseases including eczema and SLE; lipid traits like total cholesterol (TC), low-density lipoprotein cholesterol (LDL), and triglycerides (TG) (**Figure 4a**). Brain tissue associated traits have higher correlations based on estimates using fetal brain tissues (**Figure 4b**) compared to those from adult non-brain tissues. For example, Alzheimer’s disease (AD) is clustered with autoimmune-related traits in adult non-brain tissues, whereas it is correlated with psychiatric traits like bipolar disorders (BD) and ADHD in fetal brain tissues. For some other traits, their correlations in 13 fetal brain tissues were similar to those identified in adult non-brain tissues. For example, a positive correlation between ASD and ADHD was observed for both adult tissues (*R*^2^ = 0.33, p=1.4e-7) and fetal brain tissues (0.55, p=7.9e-16). Moreover, we observed correlations in different directions between fetal brain tissues and adult non-brain tissues. For example, smoking initiation (SmkInit) and asthma had a positive correlation in fetal brain tissues (0.35, p=1.1e-6) but a negative correlation in adult non-brain tissues (-0.33, p=1.2e-7). The associations identified between asthma and neuronal cells in fetal brain tissues may be supported by previous findings linking neural pathways to allergic inflammation in lungs^24,25^.

**Figure 4.**
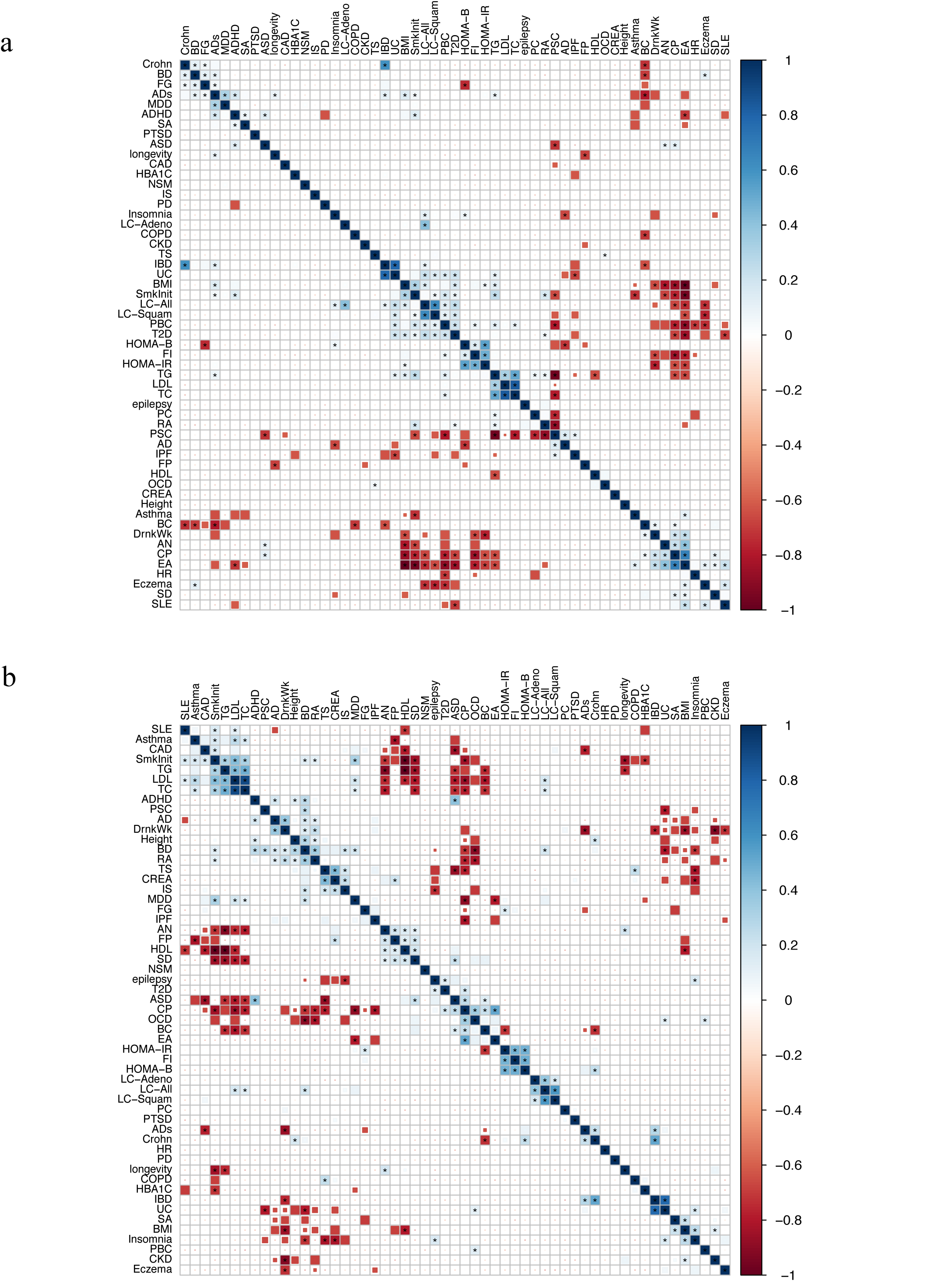
Trait-trait correlation. Different colors indicate the correlation level and the stars indicate the significant correlations after Bonferroni correction. a) Trait-trait correlation calculated from cell-disease associations in adult non-brain tissues. b) Trait-trait correlation based on cell-disease associations in fetal brain tissues.

### Breast cancer and CD8^+^ T cells

To further examine the potential utility of cWAS using specific datasets, we applied cWAS to identify cell types for breast cancer and two lung diseases. For breast cancer (BC), we used European breast cancer GWAS summary data^26^(n=228,951, n_case=122,977, n_control=105,974). In whole blood, we identified a significant negative association between GRPs of CD8^+^ T cells and BC risk (**Figure 5a**) (p=8.9e-9) using the published signature gene expression matrix LM22^12,17^.

**Figure 5.**
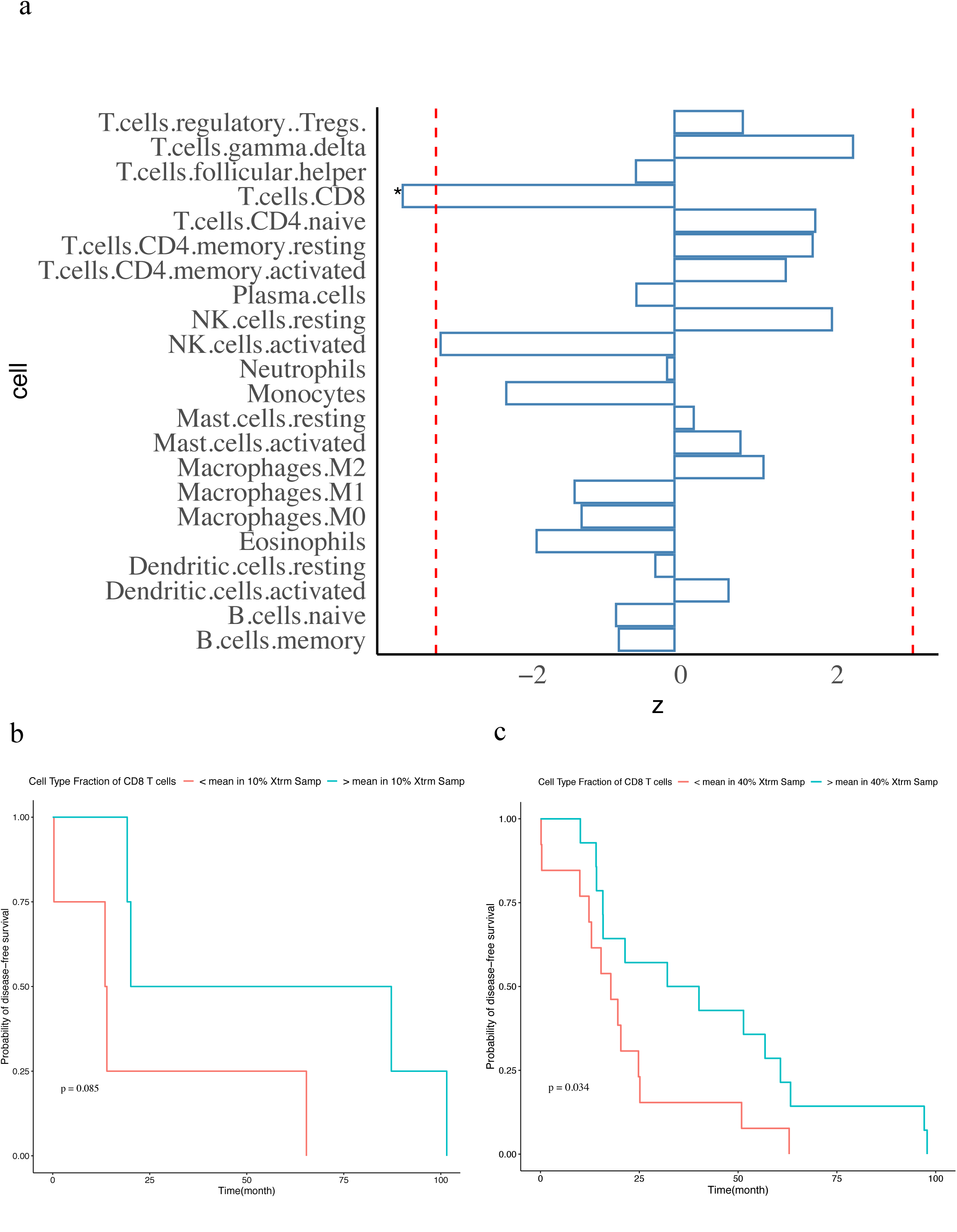
CD8^+^ T cells in breast cancer. a) cWAS results of breast cancer in whole blood. The x axis is the z score of the cell type-disease association from cWAS. Negative z scores indicate negative associations between cell type proportions and diseases. The red line indicates the significance threshold (0.05) after Bonferroni correction. The star indicates significant cell types after Bonferroni correction. b) and c) show survival analysis results in breast cancer patients of TCGA. B) Considering the white basal patients with top 10% and low 10% of genetically regulated cell type proportions of CD8^+^ T cells, and the survival patterns were compared between patients in these two groups. c) shows the results of a similar analysis in white Luminal B breast cancer patients considering patients with top 40% and bottom 40% of GRPs of CD8^+^ T cells.

To explore potential biological and clinical implications of this result, we imputed genetic-regulated cell type proportions in whole blood for subjects with European ancestry in The Cancer Genome Atlas (TCGA) project who were diagnosed with BC (TCGA-BRCA)^27^ (see **Methods**). We found that basal breast cancer patients with higher imputed CD8^+^ T cell proportions had an overall better survival (**Figure 5b**, p=0.085). Results were similar but significant (p=0.034) for luminal B breast cancer patients (**Figure 5c**). We also considered an alternative approach to evaluating cell-type specific expression patterns of BC-associated genes identified using epigenetic annotations and genetic signals (T-GEN^28^). BC-associated genes showed no significant expression enrichment in any cell type of whole blood other than a significant depletion in dividing NK T cells (fold-change=0.79, p=1.6e-8) (**S Fig. 3a**). Furthermore, BC-associated genes identified by T-GEN did not show significantly higher expression levels in T cells or any other cell types (**S Fig. 3b**).

To further validate the results, we studied BC-cell type proportion association using another cell type proportion decomposition approach^29^. In this case, the cell type proportion association result was based on the directly measured tumor tissue transcriptome data from TCGA-BRCA. We found a similar protective effect of the CD8^+^ T cell proportion (p=0.013) in basal breast cancer patients (**S Fig. 4a**), but not in luminal breast cancer patients (**S Fig. 4b, 4c**).

### Lung diseases and lung tissue

Using single cell data^30^ with better quality than HCL data to identify cell types with small proportions, we performed cWAS analysis for two lung diseases, idiopathic pulmonary fibrosis (IPF, n=24,589, n_case=4,124, n_control=20,465)^31^ and chronic obstructive pulmonary disease (COPD, n=5,346, n_case=2,812, n_control=2,534)^32^. In IPF, a higher predicted proportion of myofibroblast in lung tissue was associated with an increased risk of developing the disease (p=5.3e-4, **Figure 6a**), consistent with the accumulation of myofibroblasts observed in IPF patients^33^. We also observed a negative association of fibroblast proportions in the development of IPF (p=3.5e-2), which is consistent with aberrant fibroblast-to-myofibroblast^34^ differentiation and fibroblast degeneration and myofibroblast proliferation^35^ in IPF.

**Figure 6.**
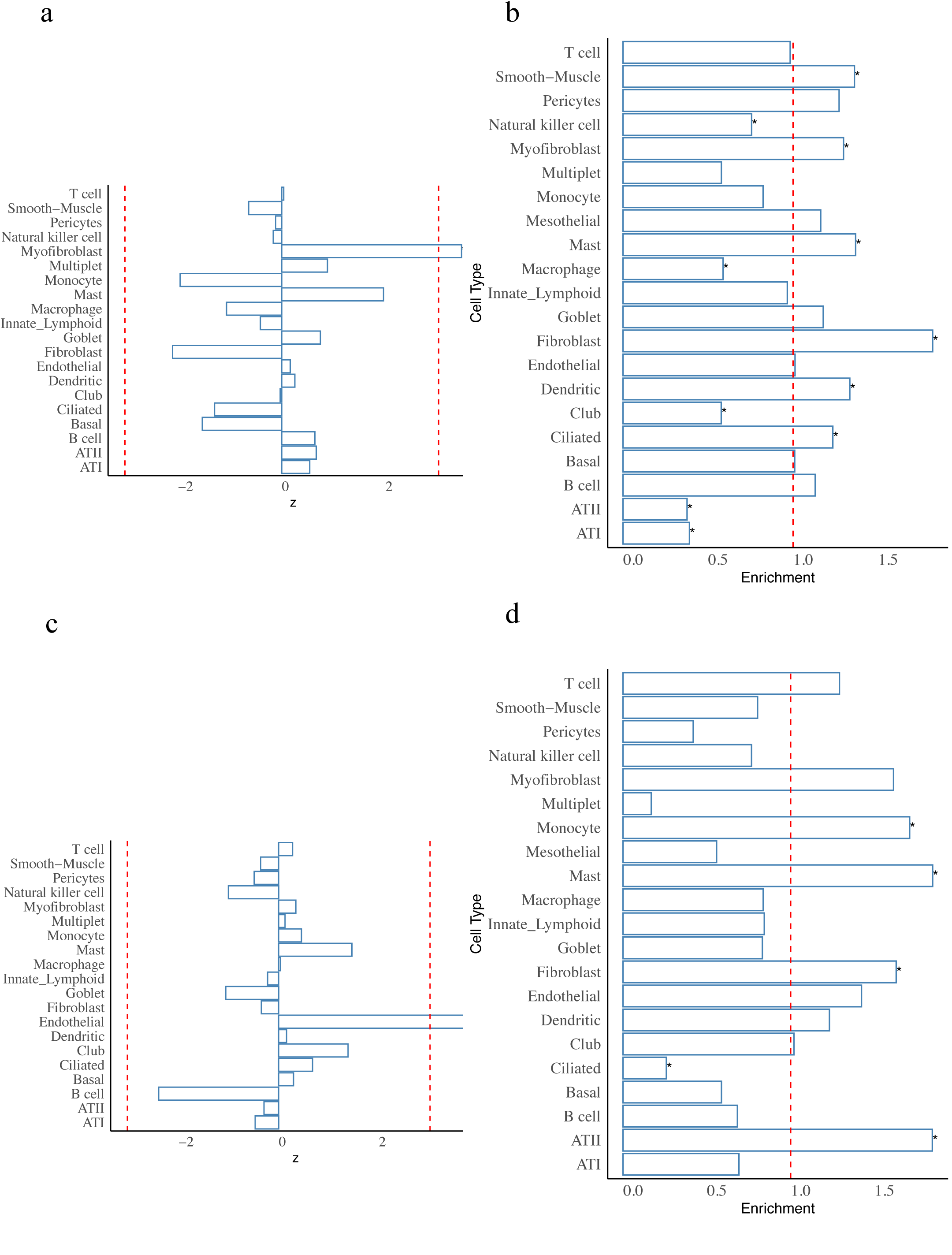
cWAS association results of IPF and COPD in lung. For a) and c), the red line indicates the significance threshold (0.05) after Bonferroni correction. For all figures, stars indicate significant cell types after Bonferroni correction. a) cWAS results of IPF in lung tissue. The x axis is the z score of the cell type-disease association from cWAS. Negative z scores indicate a negative association between cell type proportions and the disease. b) Cell-type specific expression enrichment pattern of upregulated genes in IPF patients. c) cWAS results of COPD in lung tissue. d) Cell-type specific expression enrichment pattern of upregulated genes in COPD patients.

To further evaluate cell type associations with IPF, we investigated the cell type expression pattern of IPF dysregulated genes by conventional transcriptomics analysis. Using differentially expressed genes from the published^36^ RNA-seq data of lung tissue in IPF patients (n=36) and non-disease individuals (n=19), we found that upregulated genes in IPF patients were significantly enriched in myofibroblasts (fold change=1.3, p=1.4e-3, **Figure 6b**). However, genes differentially expressed in myofibroblasts can result either from genetic effects or disease status. We further analyzed the cell type signal based on genetic information using IPF GWAS summary statistics. Applying MAGMA (implemented in FUMA, see URLs) to the IPF GWAS results (**S Fig. 5a**), we found marginal evidence of enriched genetic signals in the fibroblasts of lung tissue (p=7.5e-2). IPF-associated genes identified by T-GEN^37^ did not show any significant enrichment in any cell type of lung. Therefore, though neither was significant after Bonferroni correction, both transcriptomic and gene-set based genetic analyses suggest the importance of myofibroblasts and fibroblasts consistent with cWAS.

For COPD, cWAS found higher GRPs of endothelial cells increased disease risk (p=2.1e-4, **Figure 6c**). To further investigate the association, we applied cWAS in additional GWAS of a larger sample size with a signature matrix having more refined cell types (**Methods**). One specific endothelial cell type, vascular endothelial capillary A, was positively associated (p=3.9e-4) with COPD based on results from another GWAS (N=257,811)^38^. Upregulated genes in COPD patient lung tissue^36^ were also enriched in endothelial cells (fold change=1.4, p-value=1.4e-2) (**Figure 6d**). Similar to IPF analysis, we also investigated COPD genetic signal enrichment using MAGMA on mouse lung data (no human lung data available in FUMA, **S Table 6**). There was marginal evidence of signal enrichment in endothelial cells in the lung tissue (p=8e-2, **S Fig. 5b**) and lung vasculature (p=2.8e-2, **S Fig. 5c**). Similar to the results in IPF, T-GEN-identified genes in COPD did not show any enrichment in cell types of lung. Nevertheless, these results support the cWAS results indicating a role for endothelial cells in COPD.

We further validated the findings on IPF-myofibroblast and COPD-endothelial associations using another lung scRNA-seq dataset^30^. This recent study profiled 32 IPF patients, 18 COPD patients, and 28 controls, and we compared major cell type proportions across these three groups of samples (**S Fig. 6a**). In IPF patients, the myofibroblast cell type proportion was significantly increased (p=1.3e-3, **Figure 7a**) compared with other major cell types (**Figure 7b**). We also conducted pathway analysis on both up- and down-regulated genes in IPF myofibroblast cells (**Figure 7c**). The top enriched pathways of upregulated genes mostly function as the extracellular matrix^39^ (ECM), a network playing an important role in cell adhesion and linking glycoproteins with fibrous proteins, supporting the importance of the fibroblast-to-myofibroblast migration process in IPF. In COPD, despite low endothelial cell counts and the limited sample size in the single cell data (**S** **Fig 6b**), analysis of upregulated genes in COPD endothelial cells (**Figure 7d, S Fig 6b, 6c, 6d**) suggests the involvement of DNA-binding transcription activity and higher activity of COPD endothelial cells compared to control endothelial cells.

**Figure 7.**
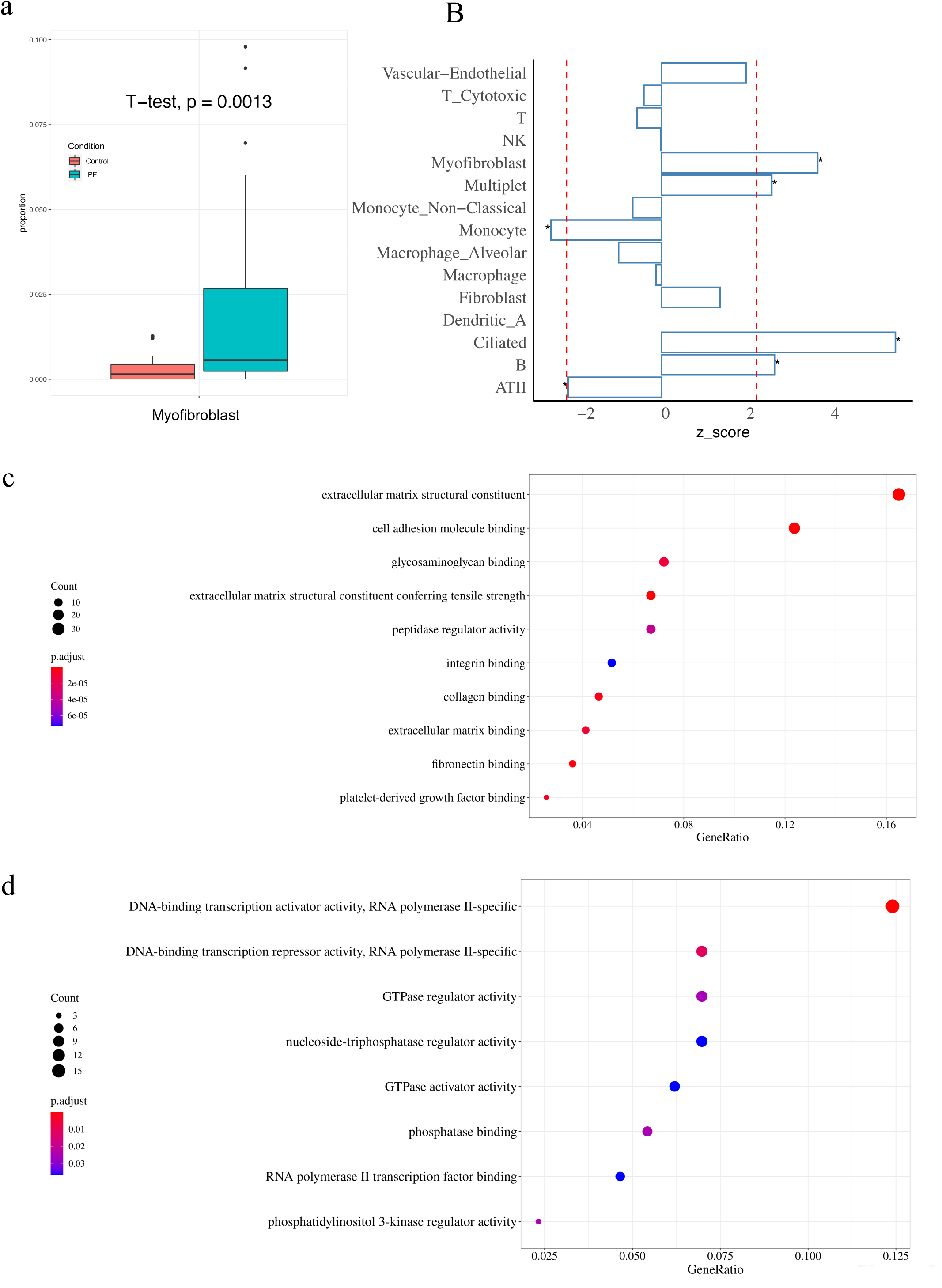
IPF myofibroblast and COPD endothelial cell type proportion validation in the separate scRNA-seq atlas. a) Boxplots of myofibroblast cell type proportions in 32 IPF patients and 28 controls. The vertical axis is the cell type proportion of myofibroblast. The IPF myofibroblast cell proportion is significantly higher than that in controls with p-value = 1.3e-3 by t-test. b) Bar plots of z scores when cell type proportions were regressed on conditions of IPF and control. The red line indicates the significance threshold (0.05) after the Bonferroni correction. The star indicates the significant cell types after Bonferroni correlation. All the cell types with z scores greater than 2 are labeled with an asterisk. Only cell types whose proportions are more than 1% are shown. Myofibroblast ranks second in these 15 major cell populations. This difference may be related to the genetically mediated regulation of cell type proportion based on the cWAS results. c) Scatterplots of Gene Set Enrichment Analysis (GSEA) results of IPF myofibroblast up-regulated genes. The dot size is the gene counts found in the pathway. The colors indicate the hypergeometric test p-values. Most top enriched pathways are related to ECM and cell adhesion. d) Scatterplots of GSEA results on COPD endothelial up-regulated genes. The dot size is the gene counts found in the pathway. The colors indicate the hypergeometric test p-values. The pathways indicate a stronger DNA-binding transcription activity.

## Discussion

Recent analyses have devoted great efforts to understand GWAS findings in traits and diseases. Several methods have been developed to link identified variants to genes based on genomic locations^2^, epigenetic annotations, or eQTL regulations^18^. At the cell type or tissue level, methods like LD score regression^40^ and FUMA^3^ either utilize annotation information or expression data to investigate the genetic enrichment pattern in cell types or tissues. Differing from previous methods, cWAS is a novel statistical framework to interpret GWAS findings in a cell type proportion manner. It helps researchers gain insights into the relationship between cell type GRPs and diseases. cWAS is complementary to cell type-disease associations identified solely through genetic association or heritability enrichment, especially when genetic signals are mediated by regulating cell type proportions. Identified disease-associated cell type proportions can potentially serve as biological markers in clinical practices to identify patients with higher genetic risk^41,42^. Applying cWAS to GWAS summary statistics from 55 traits, we found that previously genetically correlated traits also have correlated associations with GRPs of cell types. Applications of cWAS to Breast Cancer, IPF, and COPD identified cell type proportion-trait associations, which were supported by either previous findings or our analysis of other data. Specifically, a high proportion of CD8^+^ T cells was identified as protective in breast cancer development based on both transcriptome and cWAS analyses. Survival analysis using imputed GRPs of cell types also implied a protective effect of higher CD8^+^ T cell proportion in breast cancer prognosis. All these findings support the importance of CD8^+^ T cell proportions for both disease onset and prognosis in breast cancer.

We noted that transcriptome analyses of breast cancer patients have also identified the importance of CD8^+^ T cells. Utilizing breast tumor infiltration data, multiple published survival studies^43,44^ found protective effects of high CD8^+^ T cell proportions in the tumor tissue for breast cancer. In contrast to TCGA results based on observed breast tissue expression data, cWAS identifies genetically regulated cell type proportions in whole blood, which are more likely to cause the development of the disease instead of being affected by the disease status. Although the mechanisms in the prognosis and the development of breast cancer are not necessarily the same, the converging evidence from different approaches used here suggests the importance of CD8^+^ T cells in breast cancer.

We also note that previous COPD research has already implied the importance of endothelial cells^45^, which are involved in both the initiation and progression of COPD as well as other lung diseases, such as asthma and emphysema. More specifically, endothelial cells play a role in the transendothelial migration (TEM), through which neutrophils move to lung tissue and respond to the residential inflammation^46^. Additionally, endothelial apoptosis in lung initiates and contributes to the progression of COPD disease^45,47^. Previous genetic research also identified the importance of endothelial cells in COPD^38^ using ATAC-seq data and emphysema^48^. This further lends support to the involvement of endothelial cells in developing COPD.

Similar to many statistical methods, cWAS is also highly dependent on the data used, more specifically, the single cell data. The single cell data used in signature gene expression curation can affect cWAS performance, including the cell types included and the signature gene expression levels. More single cell databases with larger sample sizes, higher resolution and more comparable experiment pipelines across more tissues will aid in its further application and result interpretation. To mitigate the batch effects across various tissues, for trait-trait correlation analysis and multi-tissue association analysis, we used the HCL dataset to extract signature gene matrices since the test results will be comparable across tissues due to relatively small batch effects and the same cell labeling criteria in all tissues. For BC, we used the LM22 matrix, which was curated based on the Affymetrix microarray data, to extract the signature matrix in whole blood. In IPF and COPD, the signature matrix was curated using single cell data of lung from HCL, which consists of 23,878 cells from 20 cell types. Notably, due to randomness of obtaining samples in experiments, cell type composition in lung single cell data can be strongly biased, with 90% of the cells being immune cells. Despite this limitation, we identified a non-immune cell population in COPD. Our results based on fetal brain single cell data relied on the assumption that the genetic regulation of gene expression is the same in both adult and fetal tissues. The assumption could be violated for tissues still undergoing development in fetuses^49^. The accuracy of cWAS results could be further improved if matched genotype and cell type proportion data were available for identifying cell type proportion QTLs.

Nevertheless, future work can further expand the potential of cWAS analysis. First, considering the differentiation trajectory between cell types will further better pinpoint the associated cell types or even causal cell types, but will also limit the application of cWAS since not all differentiation trajectories are known in human tissues. Second, when analyzing specific traits across tissues to identify the most signal-enriched tissue, we found that traits like BMI and height are associated with cell types in almost all tissues, even though both BMI (p=1.4e-13) and height (p<2e-16) has the strongest signal in whole blood tissue. The results can be affected by the comparably large sample size of BMI and height GWAS as well as the complex biological processes involved in these traits. Future work can explore the potential of jointly modeling multiple traits to identify trait-specific associations with cell type proportions.

To conclude, different from bulk RNA-seq or scRNA-seq analysis comparing patients and healthy individuals, cWAS assesses the association between GRPs of cell types and diseases. In addition to typical genetic enrichment methods like MAGMA and LD score regression, cWAS provides a novel way to investigate the cell type-disease association. Both simulation and real data analyses have demonstrated the statistical power of cWAS in providing new insights in understanding the genetic etiology of human diseases from the cell type proportion perspective.

## Online Methods

### Expression imputation model training

Tissue-specific expression imputation models were trained in 44 tissues using matched individual-level RNA-seq and whole-genome sequencing data from the GTEx (v8) project. We focused on common SNPs (minor allele frequencies > 0.05) by filtering out SNPs whose allele frequencies were smaller than 0.05. RNA-seq data were adjusted for possible confounding factors, including the first five genotype principal components (PCs) and different numbers of Probabilistic Estimation of Expression Residuals (PEER) factors. Only cis-SNPs located within 1Mb from the transcription starting site of each gene were considered for training the gene expression imputation model.

Ten-fold cross-validated elastic-net models were applied to build gene expression imputation models, with the parameter *α* as 0.5 and the optimal *λ* selected via the function cv.glmnet provided in the ‘glmnet’ package. Only gene expression imputation models with FDR < 0.05 were considered in the following analysis. To make the test results more robust, we only considered those models with an imputation accuracy higher than the median level in each tissue.

### Single cell datasets preprocessing

All single-cell data used in this project were obtained from public repositories. In the trait association analysis, we obtained the tissue-specific signature matrices from the Human Cell Landscape (HCL)^22^, sequenced on the microwell-seq platform [1]. HCL provides a coherent sequencing procedure that can minimize the batch-effects to have a higher consistency, making the trait-trait correlation analysis feasible. To better utilize HCL, we manually cleaned the cell type annotation across the tissues to have a consistent cell type naming rule.

We found that the curation of signature matrices might not be representative enough if they were only based on the raw counts due to the high drop-out rate of single cell data. To alleviate this problem, We applied SAVERX^50^, a deep Bayesian autoencoder single cell imputation tool implemented with transfer learning, on the single cell expression profile to impute drop-out events before signature matrix computation. SAVERX may distinguish the dropout and real zero expression, which helps to get a more accurate cell type-specific average expression. It is common when some single cell datasets have a rare cell population. The limited cell counts make the average expression profile across cells unstable for signature matrix. Therefore, we filtered out cell types with low counts and only kept the major cell types.

In lung disease analysis, to get the signature matrix with deeper sequencing depth and accurate cell type annotations, we used control samples in the IPF cell atlas, which contains 312,928 cells from subjects without IPF and without IPF. We partitioned the lung atlas randomly to get a smaller subset with 20,000 cells. For the signature matrix with more cell types, we include all observed cell types, while the signature matrix curated for the original two GWAS summary stats only included the main 20 cell types annotated in the IPF cell atlas.

### Association analysis

After getting SNP weights 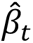 on the tissue-specific gene expression imputation models, we further combined them with published GWAS summary statistics to estimate cell-type associations with disease phenotypes. For a specific cell type *c*, we consider the association between a phenotype *Y* and its genetically regulated cell type proportions 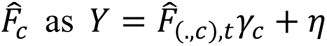. From the linear deconvolution of genetically imputed tissue-specific gene expression, we can estimate the genetically regulated cell type proportion as follows:

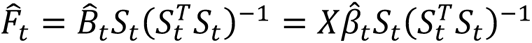

where 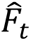 is the cell type proportion matrix in tissue *t* and *S_t_* is the expression matrix of cell-type specific signature genes.

When the individual level data are not available, we cannot obtain the cell proportions 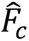. By considering the genotype-phenotype association *Y* = *X*ω + η_1_, we can indirectly estimate the coefficient *γ_c_* as follows:

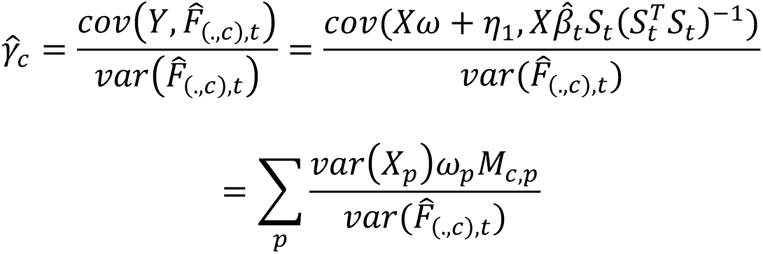

where 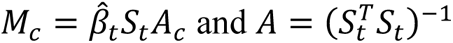.

To further get the z-score statistics for each cell type 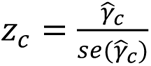, we would need to get the variance of the estimated coefficients 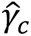. Based on simple linear regression, we can get:

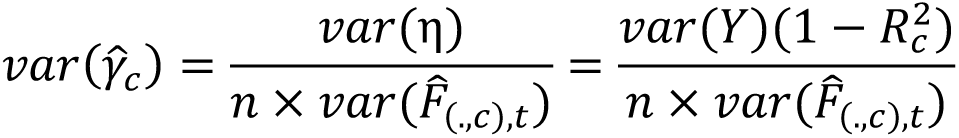

where 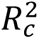 is the correlation between the phenotype *Y* and the predictor 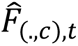. At the same time, based on the phenotype-genotype association from GWAS, we would have:

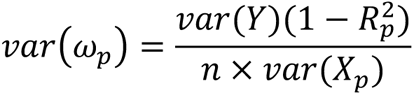

where 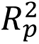 is the correlation between the phenotype *Y* and the predictor 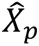. Combining the equations above, we can get the *z_c_* statistic formulated as follows:

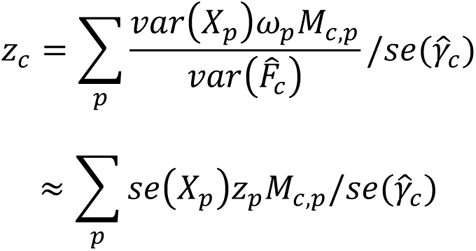

and *z_p_* is the z-score for SNP *p* for GWAS summary stats for the phenotype of interest.

### Simulation

In simulation studies, we randomly sampled 10,000 individuals from the UK Biobank dataset. Based on their genotypes of common SNPs and gene expression imputation weights trained above, we imputed their genetically regulated gene expression levels in whole blood and lung. Based on the LM22 signature matrix and simple linear regression, we imputed the cell type proportions for each sample in whole blood and used the signature matrix curated from the HCL database to get the cell proportion for lung tissue. For power analysis, we simulated phenotypes based on the imputed cell type proportion of M1 macrophages under different cis-eQTL heritability values from 0.1 to 0.09 by assuming the effect size of each cis-SNP follows the same normal distribution.

Here we defined the heritability as the phenotypic variance contributed by the imputed cell type proportion of M1 macrophages. Then we used PLINK to conduct GWAS analysis to obtain the GWAS summary results. Sex and first ten principal components of genotypes were adjusted. These GWAS summary results were used in the cWAS test to identify disease-cell type proportion association in whole blood. For the type-I error analysis, the disease phenotypes were simulated based on imputed proportions of basal cells in lung tissue. Similar to the analysis in the whole blood tissue, we obtained the GWAS summary statistics but the heritability we considered was 0.05, 0.1, and 0.5. After getting the GWAS results, we applied cWAS to identify disease-cell type proportion associations in whole blood tissue.

### Signature gene expression matrix curation

Only protein-coding genes were considered here. We selected the signature genes by differential expression (DE) analysis, i.e., Wilcoxon rank sum test, Model-based Analysis of Single-cell Transcriptomics (MAST)^51^, and ANOVA. Among these methods, MAST is a DE framework that takes cell size and drop-out rates into consideration. The Wilcoxon Rank Sum test and MAST for DE analysis were conducted by the FindMarkers() command in package Seurat (3.1.5). Bonferroni correction at *α* = 0.05 was used. When a large number of DE genes were selected, we kept the DE genes which were upregulated and differential to a single cell population. We took the intersection between significant DE genes and the GTEx-V8 genes of the corresponding tissue and included them in the signature matrix. By setting different thresholds and applying appropriate DE analysis approaches for filtering, we aimed to get the signature matrices. We computed the cell type-specific gene signature matrices by the average expression levels across cells within cell populations in the final step.

### Survival analysis in TCGA data

The imputation of the tissue-specific bulk RNA-seq expression for the TCGA-BRCA samples was based on individual germline genotypes and corresponding expression weights trained above. We followed the same procedure in the work of Huang et al. to process the germline genotypes^27^ from TCGA. Missing SNPs were not considered.

After getting the imputed tissue-level gene expression, we used linear regression to estimate the genetically regulated cell type proportions for each sample. For survival analysis, we compared the disease-free survival times between groups with a high and lower percentage of genetically predicted cell type proportion of the identified cell type. More specifically, we extracted those two groups of samples with extremely high (e.g.: top 10%) or low genetically predicted cell type proportion levels. Then, we compared disease-free survival times for the samples with the top percentage of genetically regulated cell type proportion and those with a lower percentage of genetically regulated cell type proportion levels.

### Curation of GWAS summary data

We collected GWAS summary data of 55 phenotypes, their detailed information can be found in supplementary table 2. We intentionally selected studies with most of the populations being European to reduce the bias due to population stratification. GWAS summary statistics were curated by first filtering out SNPs with minor allele frequencies less than 0.05. For datasets without rsID, we used the human genome reference build 37 to map to corresponding rsID. For datasets without Z score or P-value, we manually obtained those columns using other available information such as beta, odds ratio, and standard error. After these steps, all GWAS summary statistics contain rsID, reference and alternative alleles, Z scores, p values, and sample sizes.

For down-sampled GWAS summary stats, we considered the z scores as 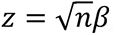, where *β* is the reported effect sizes in GWAS results and *n* is the sample size. When we reduced the sample size of the GWAS summary stats, we consider the 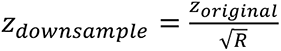 where *R* is the ratio of the sample size in the original GWAS summary stats over the sample size in the down sampled GWAS summary stats. We considered *R* = 2, 3, 4, 5 in our study here.

### MAGMA

To generate annotations, gene location files using the human genome reference panel build 37 were downloaded from the MAGMA software website as the input of the gene location file. The SNP location file was generated by extracting SNPs from curated GWAS summary data and mapping to genome locations using build 37. Annotations were generated with the command: magma --annotate --snp-loc [SNPLOC_FILE] --gene-loc [GENELOC_FILE] --out [ANNOT_PREFIX].

Next, gene analysis was performed for each phenotype using the annotation files generated from the previous step. European panels of the 1000 Genomes phase 3 data downloaded from the MAGMA software website were used as the reference. The following command was used to generate gene analysis results: magma --bfile [REFDATA] --gene-annot [ANNOT_PREFIX].genes.annot --pval [PVAL_FILE] ncol=N snp-id=SNP pval=P --out [GENE_PREFIX]. Finally, 161 processed single cell expression datasets provided by MAGMA were downloaded. To avoid duplicated cell types in multiple datasets from the same data resource, we manually selected 60 datasets (**S Table 6**) for the following analysis. MAGMA gene-property analyses (v1.07) were performed using the output of gene analysis and gene expression datasets processed as described above using the following command: magma --gene-results [GENE_PREFIX].genes.raw --gene-covar [SCDATA] --model condition-hide=Average direction=greater --out [OUT_PREFIX]. Bonferroni corrections were performed per dataset during the gene-property analyses to obtain significantly associated cell types.

### Trait-cell type association analysis across tissues

Using the signature gene matrix processed from the HCL database, we applied cWAS to obtain cell type association results for each trait across different tissues. To investigate trait-trait correlation, we considered the test-statistics (z scores) of all cell types in a trait as the representation vector of the trait. Then for any two traits, we computed the Pearson correlation between their two z score vectors and the corresponding p-value to quantify the similarity between these two traits with respect to cell type associations. Similarly, to consider the correlation of the effects of a shared cell type between tissues, for a specific cell type, we would treat its association z scores with all traits in one tissue as a vector *v*_1_. We then put its association z scores with all traits in a second tissue as a vector *v*_2_. To study the tissue-tissue correlation for the shared cell type effects, we calculated the Pearson correlation between *v*_1_ and *v*_2_.

In the across-tissue analysis, for each trait, we firstly identified the significant cell type associations after the Bonferroni correction in each tissue. Then across all tissues, we identified the most significant cell type association signals.

### Differentially expressed genes from bulk and cell type enrichment analysis

Differentially expressed (DE) genes in IPF and COPD patients were downloaded from previous publications^36,52^. We curated the cell-type specific gene expression matrix in lung tissue using the published single cell data. Then for each gene, we identified the cell type with the highest expression. Then for each cell type, we analyzed the enrichment pattern of upregulated genes in patients compared to other genes in the cell type. The binomial test was used to test the significance level of the enrichment pattern and then Bonferroni correction was further applied to select the significant cell types.

The Gene Set Enrichment Analysis was conducted by the gseGO() command in package clusterProfiler (3.14.3). All the parameters were set to the defaults values, where Benjamin–Hochberg correction at *±* = 0.05 was used as the cutoff.

### URLs

Human cell landscape: http://bis.zju.edu.cn/HCL/

GTEx data: https://gtexportal.org/home/

Roadmap Epigenomics project: https://egg2.wustl.edu/roadmap/web_portal/

BC summary stats: http://bcac.ccge.medschl.cam.ac.uk/

COPD summary stats: https://pubmed.ncbi.nlm.nih.gov/24621683/;

https://pubmed.ncbi.nlm.nih.gov/30804561/

IPF summary stats: https://github.com/genomicsITER/PFgenetics

MAGMA: https://github.com/Kyoko-wtnb/FUMA_scRNA_data

## Supporting information

All supplementary table

## Data Availability

Result files and corresponding code will be available upon acceptance of the manuscript.

## Acknowledgements

Part of the this work was supported by funding from NIH grant R01GM134005 and NSF grant DMS1902903 to H.Z., NHLBI grants R01HL127349, R01HL141852, U01HL145567, UH2HL123886 to N.L. and a generous gift from Three Lakes Partners to N.K. M.H.C. was supported by R01HL135142, R01 HL137927, R01 HL089856, R01 HL147148. The research was partially supported by the NIHR Leicester Biomedical Research Centre; the views expressed are those of the authors and not necessarily those of the NHS, the NIHR or the Department of Health.

## Conflicts of Interest

N.K. served as a consultant to Boehringer Ingelheim, Third Rock, Pliant, Samumed, NuMedii, Theravance, LifeMax, Three Lake Partners, Optikira, Astra Zeneca, Augmanity over the last 3 years, reports Equity in Pliant and a grant from Veracyte and Boehringer Ingelheim and non-financial support from MiRagen and Astra Zeneca. N.K. as IP on novel biomarkers and therapeutics in IPF licensed to Biotech. M.H.C. has received grant support from GSK and Bayer, consulting or speaking fees from Genentech, AstraZeneca, and Illumina. L.V.M. holds a GSK/British Lung Foundation Chair in Respiratory Research.

## Author Contributions

W.L., W.D. and M.C. developed the statistical framework. W.L., W.D. and M.C. performed statistical analysis.

Z.D. assisted in analyzing single cell data.

B.Z. performed pathway enrichment analysis.

Y.Z. assisted in analyzing GTEx data.

D.T. analyzed the TCGA infiltration data and cell type proportional analysis.

M.S., L.V.W., M.H.C., N.K., and H.Z. advised on the biology of lung diseases.

H.Z. advised on statistical and genetic analysis.

W.L. implemented the software.

W.L., W.D. and M.C. wrote the manuscript.

All authors contributed to manuscript editing and approved the manuscript.

---

**S1 Figure.**
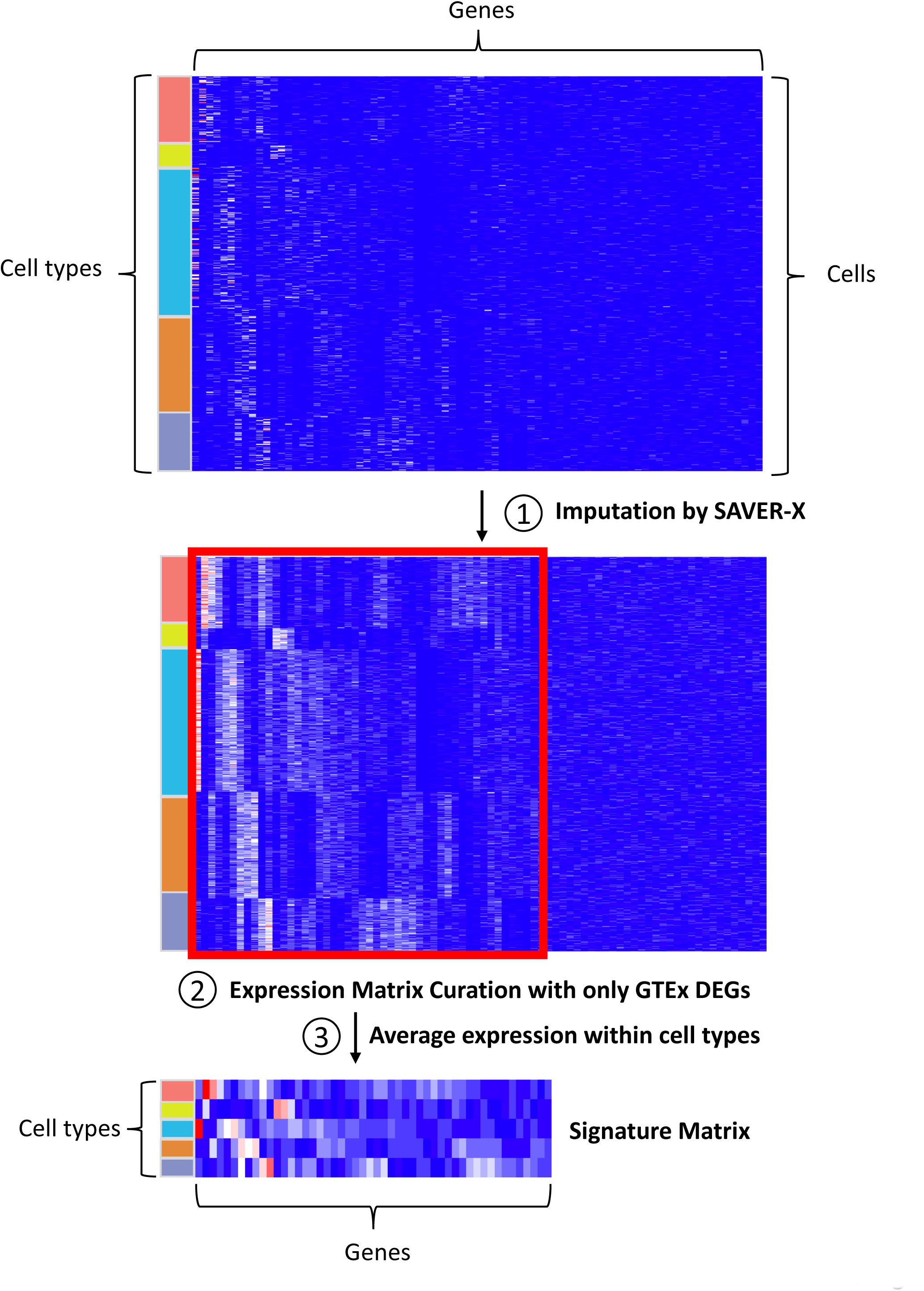
The workflow of curating gene expression signature matrix in each tissue. Single cell data across multiple cell types in tissue is firstly imputed by SAVER-X and then significant differentially expressed (DE) genes are identified based on cell-type level DE analysis. Finally, for those identified DE genes, their average gene expression levels are computed within each cell type.

**S2 Figure.**
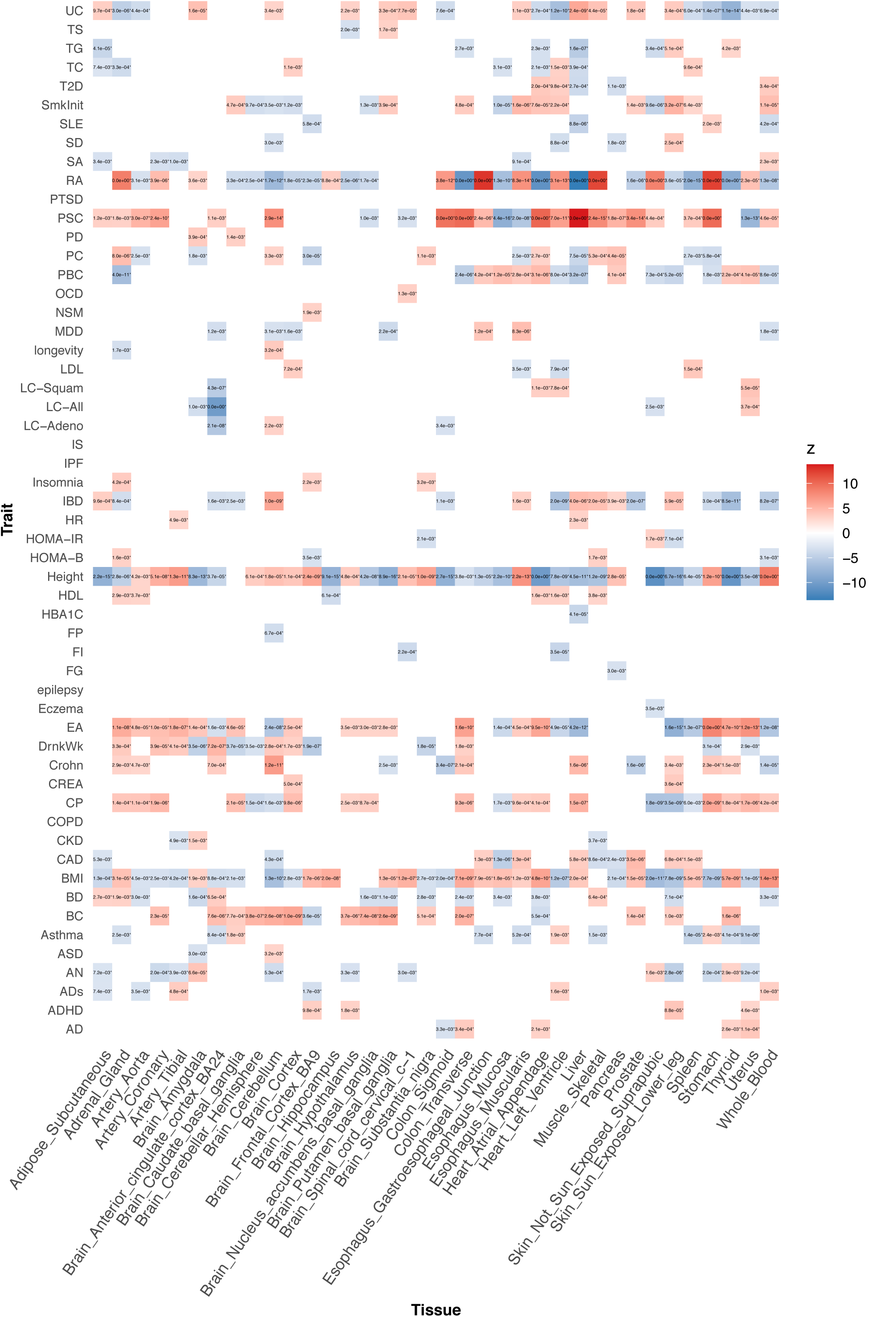
The cell type-trait associations across 55 traits identified by cWAS. In 36 tissues, the significant/most-significant associated cell types are shown in the figure. Blue colors indicate the negative correlations between traits and the corresponding associated cell type proportion while red colors indicate the positive correlations.

**S3 Figure.**
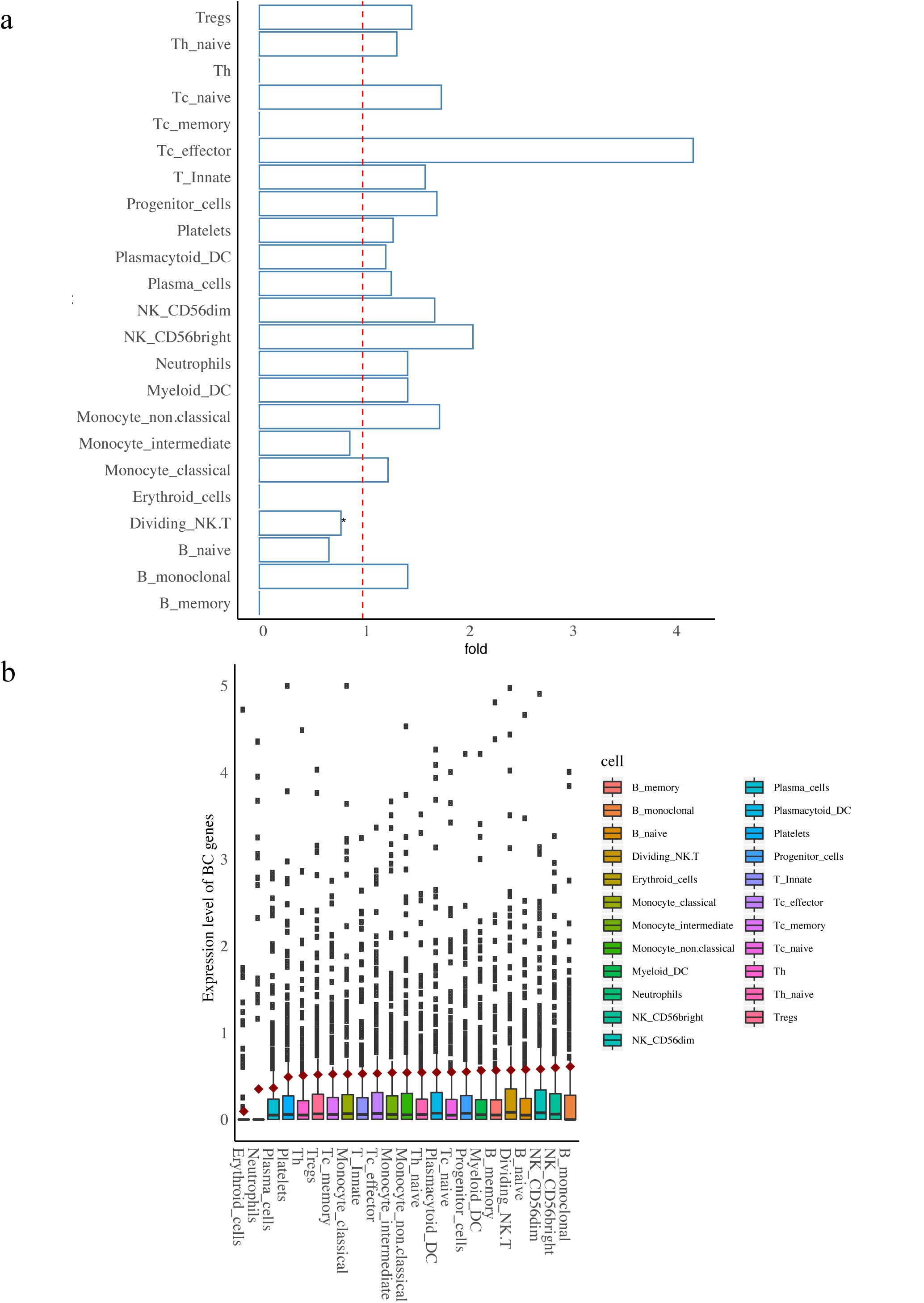
Cell type expression pattern of breast cancer (BC)-associated gene identified by TWAS analysis. a) As in previous figures, the star indicates the significant cell types after Bonferroni correction in whole blood. The fold indicates (x axis) the enrichment level of BC-associated genes in those genes with high expression specificity in the corresponding cell type. b) This figure shows the expression level of identified BC-associated genes in different cell types of whole blood.

**S4 Figure.**
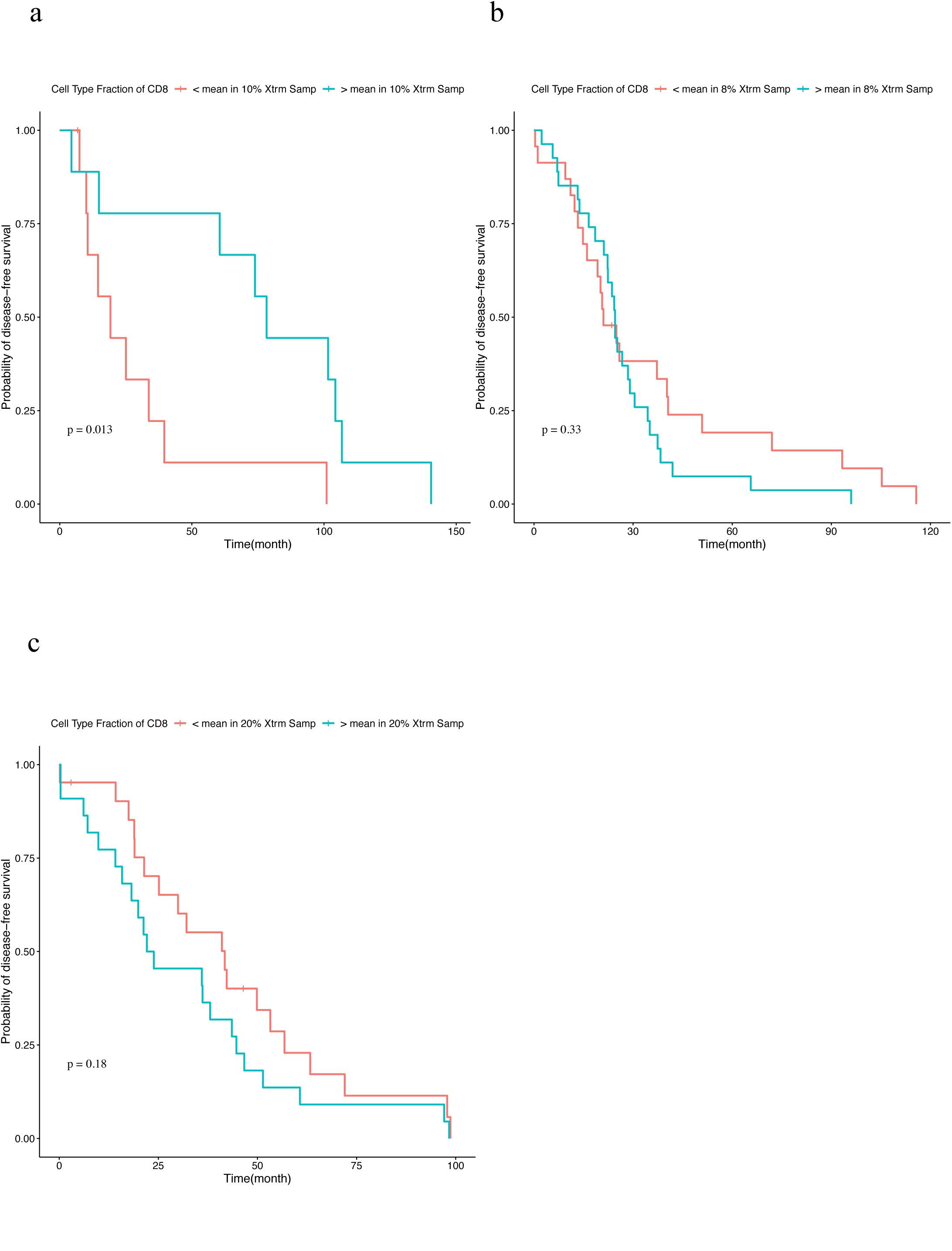
Survival analysis results of breast cancer patients in TCGA. Here we consider the cell type proportion estimated based on the assayed expression level in tumor tissues from TCGA. a) In patients of European ancestry with basal breast cancer, only patients with the top 10% and bottom 10% proportion of CD8^+^ T cells were considered. b) In Luminal A patients with European ancestry, only patients with top 8% and bottom 8% proportion of CD8^+^ T cells were considered. c) In luminal B patients of European ancestry, only patients with the top 20% and bottom 20% proportion of CD8^+^ T cells were considered.

**S5 Figure.**
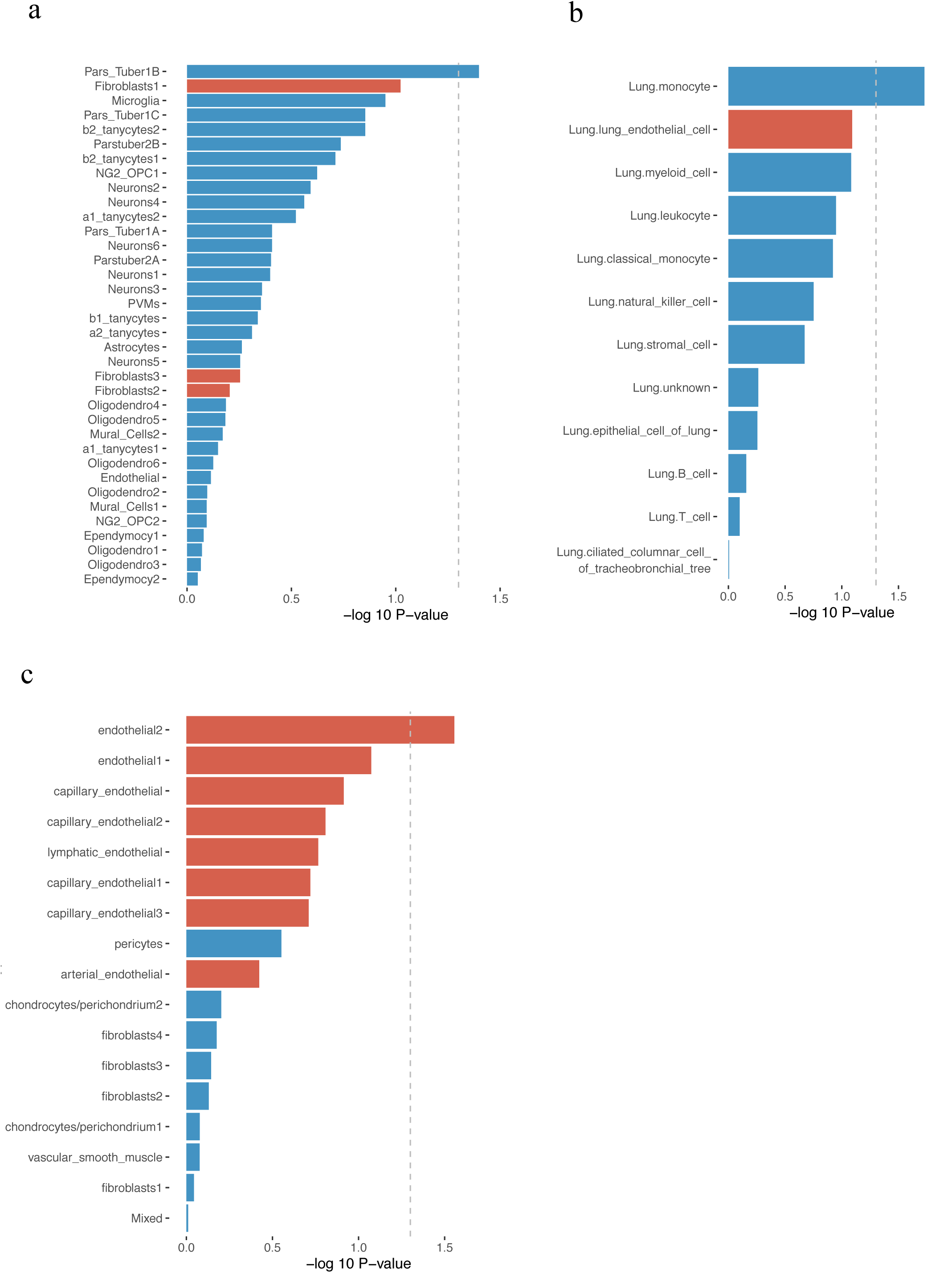
MAGMA analysis results of IPF and COPD GWAS summary stats. In all figures, the vertical dash line indicates the significance threshold after Bonferroni correction. The red bars indicate the corresponding cell types of interest in IPF and COPD. a) Bar plots of MAGMA cell type association results between IPF and all cell types of the MAGMA processed GSE93374_Mouse_Arc_ME_level2 dataset^53^. Fibroblast-related cell types are highlighted in red. The grey dashed line represents the 0.05 significance level. b) Bar plots of MAGMA cell type association results between COPD and cell types from lung tissue of the MAGMA processed TabulaMuris_FACS_all dataset^54^. Endothelial-related cell types are highlighted in red. The grey dashed line represents the 0.05 significance level. c) Bar plots of MAGMA cell type association results between COPD and all cell types from the MAGMA processed GSE99235_Mouse_Lung_Vascular dataset. Endothelial-related cell types are highlighted in red. The grey dashed line represents the 0.05 significance level.

**S6 Figure.**
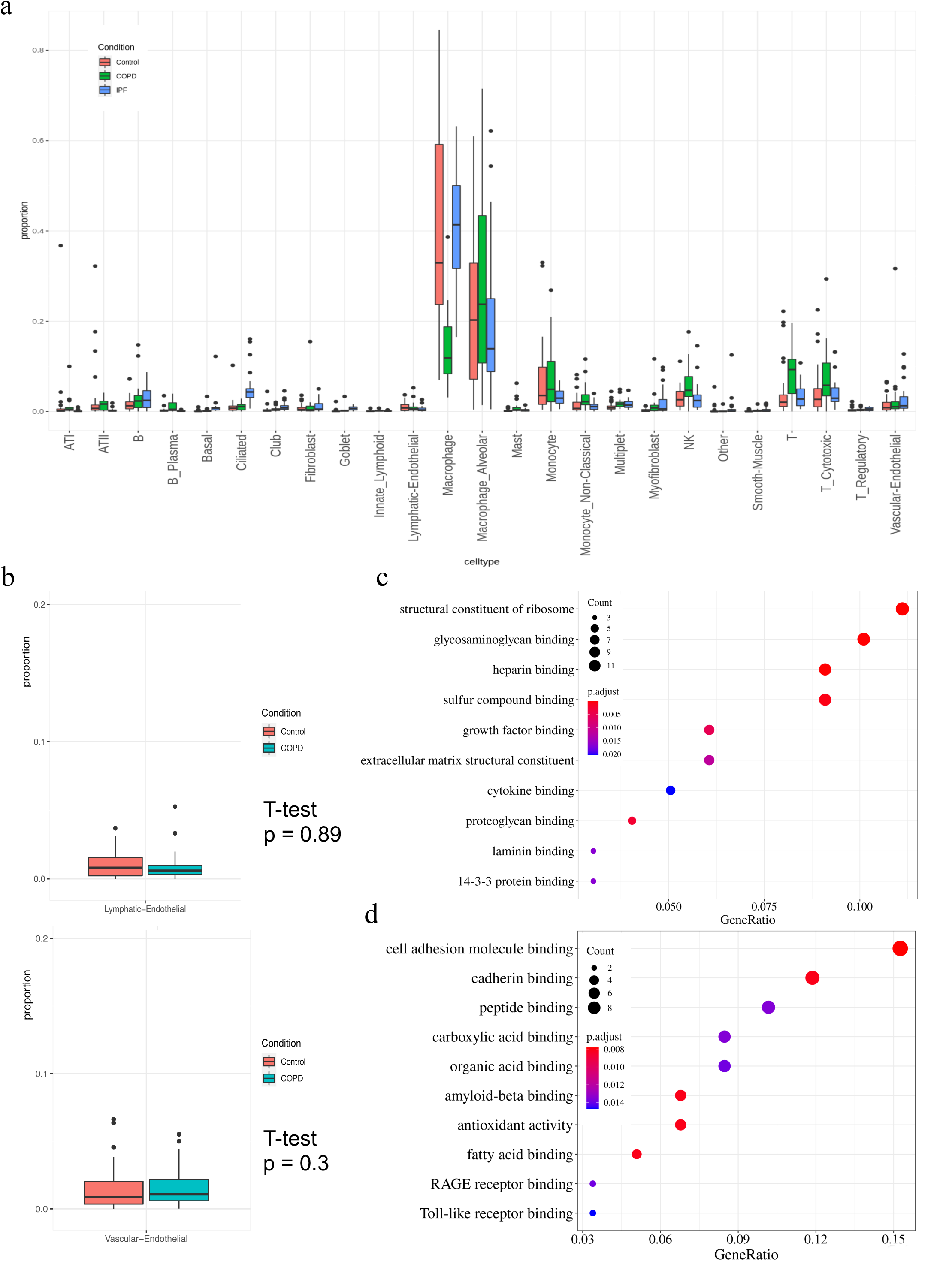
IPF myofibroblast and COPD endothelial cell type proportion validation in the separate scRNA-seq atlas. a) Boxplots of cell-type proportions comparison across IPF, COPD, and controls in lung tissue. The horizontal axis represents the major cell types. The vertical axis is the cell type proportions. The immune cells are the majority of the data. The cell type proportion has a non-negligible variance across different conditions. b) Boxplots of two endothelial subtype proportions comparison between COPD and controls. The vertical axis represents cell-type proportions. To compare the cell type proportion distributions between COPD and controls, we conducted a t-test which was not significant. However, the direction is consistent with cWAS finding for vascular endothelial. We still consider these results inconclusive due to the low endothelial cell counts. c) Dot plots of GSEA on IPF myofibroblast down-regulated genes. The dot size is the gene counts found in the pathway. The colors indicate the hypergeometric test p-values. d) Dot plots of GSEA on COPD endothelial down-regulated genes. The dot size is the gene counts found in the pathway. The colors indicate the hypergeometric test p-values.

S Table 1. Statistical power and type I error of FUMA in the simulation study

S Table 2. HCL tissues used in the analysis

S Table 3. GWAS summary statistics for 55 traits used in the trait-trait correlation analysis

S Table 4. Shared cell types (mainly immune cells) in all tissues

S Table 5. cWAS test results in all HCL tissues for 55 traits

S Table 6. scRNA-seq data sets used for MAGMA analysis

## Notes

### Author Declarations

This manuscript is a mainly methodology paper. The data used in this manuscript is all public available. The research has no procedure related to collecting human subject data.

